# Quantifying cortical lesions in large legacy multiple sclerosis clinical trial MRI datasets using multi-contrast post-processing and deep learning

**DOI:** 10.1101/2025.04.08.25325461

**Authors:** Michael G. Dwyer, Niels Bergsland, Alexander Bartnik, Dejan Jakimovski, Samantha Noteboom, Menno M. Schoonheim, Martijn D. Steenwijk, Jinglan Pei, David Clayton, Robert Zivadinov

## Abstract

Multiple sclerosis (MS) is a chronic neurological disease affecting both white and gray matter of the central nervous system. Despite the well-established history of gray matter involvement in MS, cortical lesions are almost never evaluated in clinical trials because of limitations in the feasibility of magnetic resonance imaging (MRI) to visualize them. Recently, a number of post-processing methods, including synthetic contrasts and artificial intelligence (AI)-based approaches, have shown potential for enhancing cortical lesion detection on conventional MRI data. These methods have the potential to reanalyze existing clinical trial data to answer key mechanistic questions about both MS development and about treatment effects. Therefore, we evaluated three of the most promising of them – FLAIR^2^, T1/T2 ratio, and AI-DIR – and introduced a new combined contrast called multi-modal cortical lesion enhanced (MMCLE). We also harnessed transformer-based semantic segmentation to improve automated detection and delineation of these lesions. Using the data from the large, multicenter, phase 3 ORATORIO trial, we confirmed that cortical lesions can be clearly visualized and quantified with these methods. At baseline, we detected 14.8+/-20.72 lesions per participant, 86.0% true positive rate, 8.4% false positive rate across subjects for blinded MMCLE, using simultaneous review of all contrasts as the reference. Using deep learning, we also confirmed that the simultaneous use of multiple contrasts improves quantification.

## Introduction

Multiple sclerosis (MS) is a chronic, immune-mediated disorder of the central nervous system (CNS), and is the most common disabling neurological disease of young adults. For much of its history, research and clinical care have focused on the white matter, in which focal demyelinating lesions are a hallmark of the disease.^1^ We now have many therapies that are able to almost completely arrest the incidence of new white matter lesions in individuals with MS. Disappointingly, though, they have not had the same impact on clinical progression.^2^ Over the last few decades, it has become increasingly clear that grey matter is involved from the earliest disease stages,^3^ that its pathology is not just secondary to white matter damage,^4,5^ and that targeting it may require new approaches.^6^

In particular, modern histopathological staining methods have revealed extensive cortical lesions in MS, including large subpial lesions extending across multiple gyri, leukocortical lesions straddling the boundary between gray and white matter, and smaller intracortical lesions located entirely within the gray matter.^7–9^ These lesions have been shown to occur in early disease stages across all phenotypes,^10,11^ and ultra-high-field (7T) imaging has indicated that they are present in up to 96% of people with MS.^12^ They are also highly specific to MS,^13^ and have been part of the international diagnostic criteria since the 2017 revision.^14^ From a clinical perspective, cortical lesions are strongly associated with clinical disability and cognitive impairment,^15–20^ and may have more prognostic value than white matter lesions for disability and disease course.^21^

Given this pathology and clinical relevance, there is a pressing need for in vivo methods capable of imaging cortical lesions to better understand their dynamics and response to therapy. Observationally, they have been investigated with 7T MRI, the current de facto standard, which provides excellent resolution and contrast to identify these subtle but often large areas of tissue damage.^22,23^ However, 7T availability is highly limited and infeasible in large clinical trials or most clinical practices. Advanced techniques for 1.5T and 3T have also been developed, including double inversion recovery (DIR),^24^ phase-sensitive inversion recovery (PSIR),^25^ and inversion recovery susceptibility-weighted imaging with enhanced T2 weighting (IR-SWIET),^26^ but these also pose many feasibility barriers, such as lengthened scan times, lack of standards (preventing harmonization of acqusitions across different scanner platforms), and lack of commercial sequences. Notably, when cortical lesions were included in the MS diagnostic criteria, the panel explicitly noted that their use would be greatly limited by the capabilities of clinical MRI.^14^ Because of these issues, one of the most promising sources of large-scale, high-quality evidence about disease mechanisms and treatment responses – MRI data from randomized, double-blinded Phase III trials – has not yet provided any clear information about cortical lesions.

However, in contrast to prospective approaches with their attendant feasibility issues for large trials, three recent methods have emerged for retrospectively detecting cortical lesions on available conventional images. All three are based on the idea that there is vital latent information in the relationships *between* acquired images that is invisible on the individual images themselves. First, a simple multiplicative combination of FLAIR and T2 images called FLAIR^2^ was shown to improve cortical/leukocortical sensitivity without sacrificing reliability at 3T.^27^ Second, the ratio between 3T T1- and T2-weighted images (T1/T2 ratio) was validated against 7T and showed comparable sensitivity to 3T IR-SWIET, albeit in a small population.^28^ Third, a deep learning convolutional network approach was shown to be capable of fusing T1, T2, PD, and FLAIR images into a synthetic DIR (AI-DIR) with 92% ICC against true DIR in a multi-center study.^29,30^

Against this background, we hypothesized that combined application of these novel contrasts along with AI-based detection would provide a strong approach for rapid and reliable quantification of cortical lesions in legacy clinical trial datasets, unlocking this currently-untapped source of experimental data. To test this, we evaluated FLAIR^2^, T1/T2 ratio, and AI-DIR, as well as a novel combination of FLAIR^2^ and AI-DIR called multi-modal cortical lesion enhanced (MMCLE), in a legacy clinical trial dataset – the ORATORIO trial of ocrelizumab in progressive MS.^31^ To evaluate the relative value of each contrast, we first used all four simultaneously to produce reference segmentations. Then, we assessed contrast to noise ratio (CNR) and blinded rater sensitivity/specificity on each contrast individually. To evaluate more subtle distinctions, we also trained individual deep-learning AI models on each contrast to compare detectability and reliability, as well as a final combined model to evaluate the potential value of simultaneous use of multiple contrasts. Finally, we explored potential methodological improvements for longitudinal cortical lesion activity detection and delineation.

## Results

The 80 patients from ORATORIO used for the analysis in this study had a mean age of 46.6 years, with a median EDSS of 4.5, and a mean disease duration of 7.0 years. All had primary progressive MS, per the original trial inclusion criteria. MRI scans were from 21 different scanner models (35% GE, 28.75% Siemens, 27.5% Philips, 7.5% Toshiba, 1.25% Hitachi; 81.0% 1.5T, 19.0% 3T). Additional demographic, clinical, and MRI variables are reported in Table 1. On simultaneous combined contrast analysis of baseline images, we identified a total of 1182 cortical lesions, with a mean of 14.8 per subject, as shown in Table 1. On the full trial complement of 732 subjects (used for training a final, deployable AI segmentation model and for longitudinal training), we identified 10,366 cortical lesions, with a mean of 14.4 per subject, and 193 new or enlarging cortical lesions, with a mean of 0.3 (sd=1.0, min-max=0-9) per subject. Demographic and clinical data for the full participant set has been reported previously.^32^

**Table 1.** Demographic, clinical, and MRI characteristics of the sample participants. IQR – interquartile range. ^a^T2 and contrast-enhancing lesion volumes derived from original analyses reported in the primary trial manuscript.

| **<u>Characteristic</u>** | **Analysis cohort**<br>**(n=80)** |
| --- | --- |
| **Disease course, n (%)** | PPMS, 80 (100%) |
| **Age, mean (SD.) years** | 46.6 (7.1) |
| **Sex** | 38 female (47.5%) |
| **Disease duration, mean (SD) years** | 7.02 (4.31) |
| **EDSS, median [IQR]** | 4.5 [3.5-6.0] |
| **T2 lesion volume, mean (SD) ml<sup>a</sup>** | 13.19 (14.79) |
| **Contrast enhancing lesion volume, mean (SD) ml<sup>a</sup>** | 0.50 (1.13) |
| **Baseline cortical lesion number, mean (SD, min- max)** | 14.78 (20.72, 0-121) |
| **Baseline cortical lesion volume, mean (SD, min- max) ml** | 0.80 (1.52, 0-9.93) |

### MMCLE builds on prior methods to provide high contrast to noise for lesion detection

Representative lesion samples on each cortical lesion sensitive contrast (MMCLE, FLAIR^2^, AI-DIR, T1/T2 ratio), as well as conventional FLAIR and T2 for comparison, are shown in Figure 1. Table 2 reports the contrast characteristics for all observed cortical lesions on each relevant cortical-lesion-sensitive MRI contrast type, as well as on conventional contrasts. The highest CNR was found on MMCLE, followed by FLAIR^2^, although both had higher standard deviations than AI-DIR and T1/T2 ratio.

**Figure 1.**
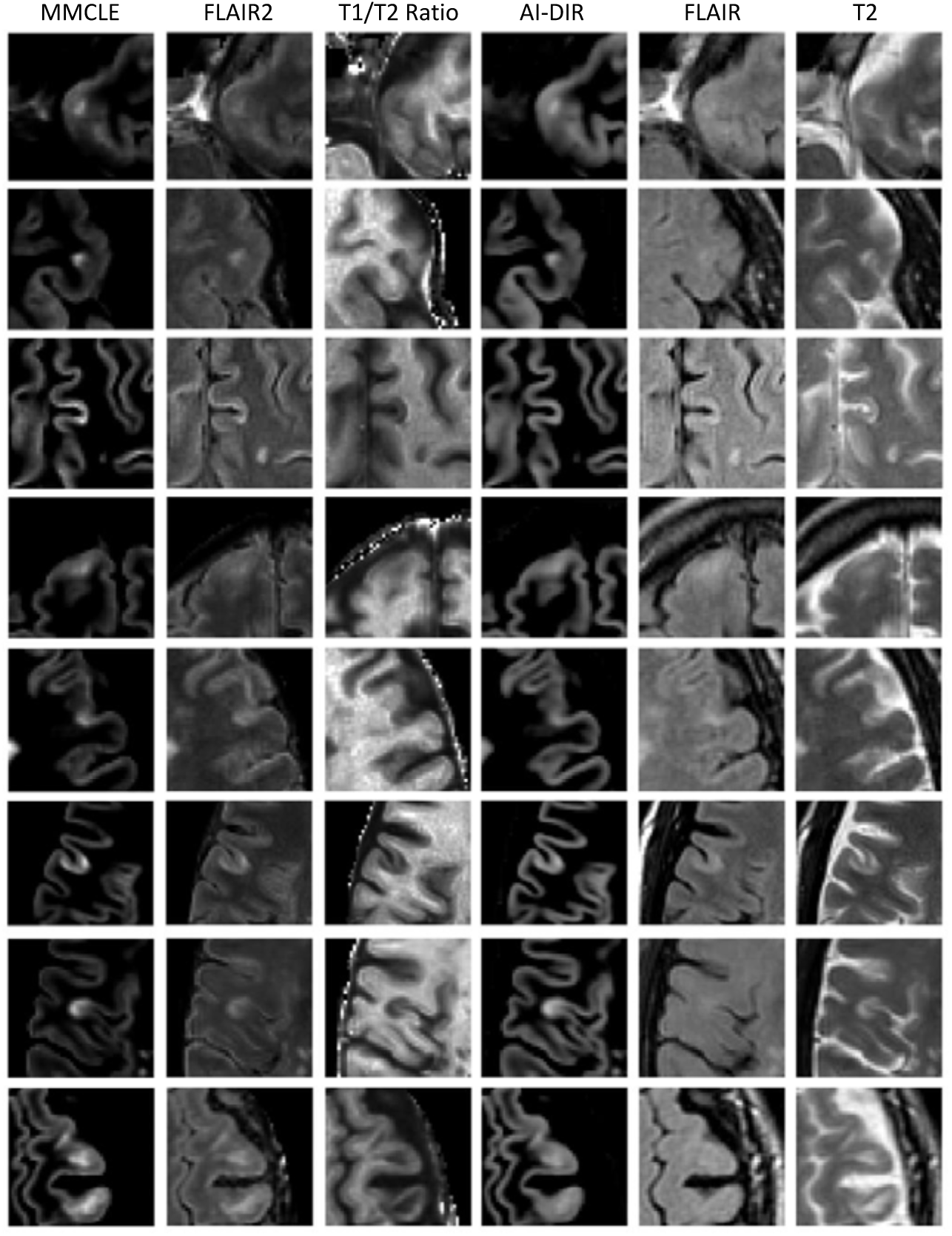
Representative examples of individual cortical lesions as seen during analysis on the various image contrasts, including both leukocortical and subpial lesions. Cortical lesion visibility varies substantially between contrasts and is not consistent across lesions. Cortical lesions were most frequently detectable on MMCLE. They were occasionally detectable post-hoc on conventional FLAIR or T2 (rows 3, 5, 7), and in rarer instances detectable a priori (row 6).

**Table 2.** Lesion contrast characteristics on each image type for the 80 selected patients. Values reported are mean and standard deviation (between subjects) of contrast to noise measured between cortical lesions relevant tissue type (columns) for each of the relevant image contrasts (rows). Rows are listed in descending order of CNR relative to NABT. MMCLE – multi-modal cortical lesion enhanced; AI-DIR – artificial intelligence double inversion recovery; CNR – contrast to noise ratio; NABT – normal appearing brain tissue; NAGM – normal appearing gray matter; NAWM – normal appearing white matter.

| Modality | CNR relative to NABT | CNR relative to NAGM | CNR relative to NAWM |
| --- | --- | --- | --- |
| **MMCLE** | 2.82 (0.83) | 2.70 (0.99) | 4.13 (1.12) |
| **FLAIR<sup>2</sup>** | 2.50 (0.75) | 1.94 (0.67) | 3.93 (1.21) |
| **AI-DIR** | 1.62 (0.32) | 1.49 (0.45) | 2.38 (0.52) |
| **T2** | 1.50 (0.56) | 0.92 (0.50) | 3.34 (1.10) |
| **FLAIR** | 1.41 (0.47) | 1.17 (0.35) | 2.07 (0.75) |
| **T1/T2 ratio** | 1.07 (0.27) | 0.53 (0.27) | 2.32 (0.49) |

### All cortical-lesion-sensitive contrasts substantially outperform conventional imaging in blinded per-contrast analysis

Results of blinded per-contrast review by 3 independent raters are presented in Table 3. The highest kappa for lesion detection (averaged across raters) was for MMCLE, at 0.78, followed by T1/T2 ratio at 0.60. MMCLE also had the highest true positive rate (TPR), resulting in average positive identification of 86.0% of lesions, followed again by T1/T2 ratio at 74.6%. However, T1/T2 ratio also resulted in nearly double the false positive rate (FPR), averaging 14.9% compared to 8.4%. Inter-rater agreement was highest on MMCLE, followed by AI-DIR.

**Table 3.**
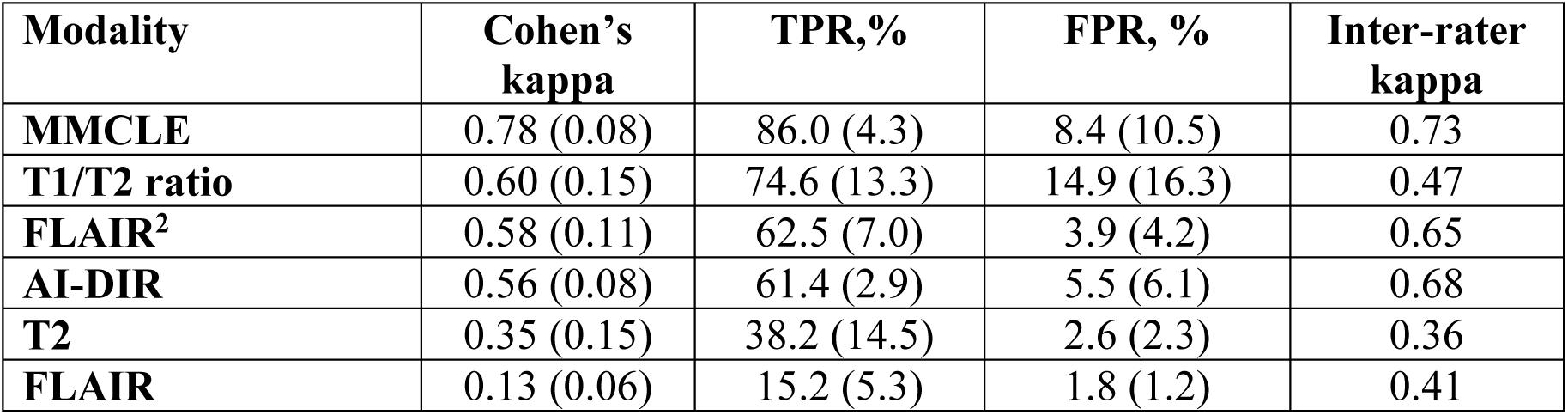
Blinded rater review of lesion detectability by contrast, as assessed by random presentation of lesions and matched non-lesions with random interleaving of contrasts. The first three columns compare raters against ground truth (consensus unblinded review of all contrasts simultaneously) and are reported as mean (SD) across raters. The fourth column compares raters between each other and is reported as Fleiss’ kappa. Rows are listed in descending order of Cohen’s kappa.

| Modality | Cohen's kappa | TPR,% | FPR, % | Inter-rater kappa |
| --- | --- | --- | --- | --- |
| **MMCLE** | 0.78 (0.08) | 86.0 (4.3) | 8.4 (10.5) | 0.73 |
| **T1/T2 ratio** | 0.60 (0.15) | 74.6 (13.3) | 14.9 (16.3) | 0.47 |
| **FLAIR<sup>2</sup>** | 0.58 (0.11) | 62.5 (7.0) | 3.9 (4.2) | 0.65 |
| **AI-DIR** | 0.56 (0.08) | 61.4 (2.9) | 5.5 (6.1) | 0.68 |
| **T2** | 0.35 (0.15) | 38.2 (14.5) | 2.6 (2.3) | 0.36 |
| **FLAIR** | 0.13 (0.06) | 15.2 (5.3) | 1.8 (1.2) | 0.41 |

**Table 4.**
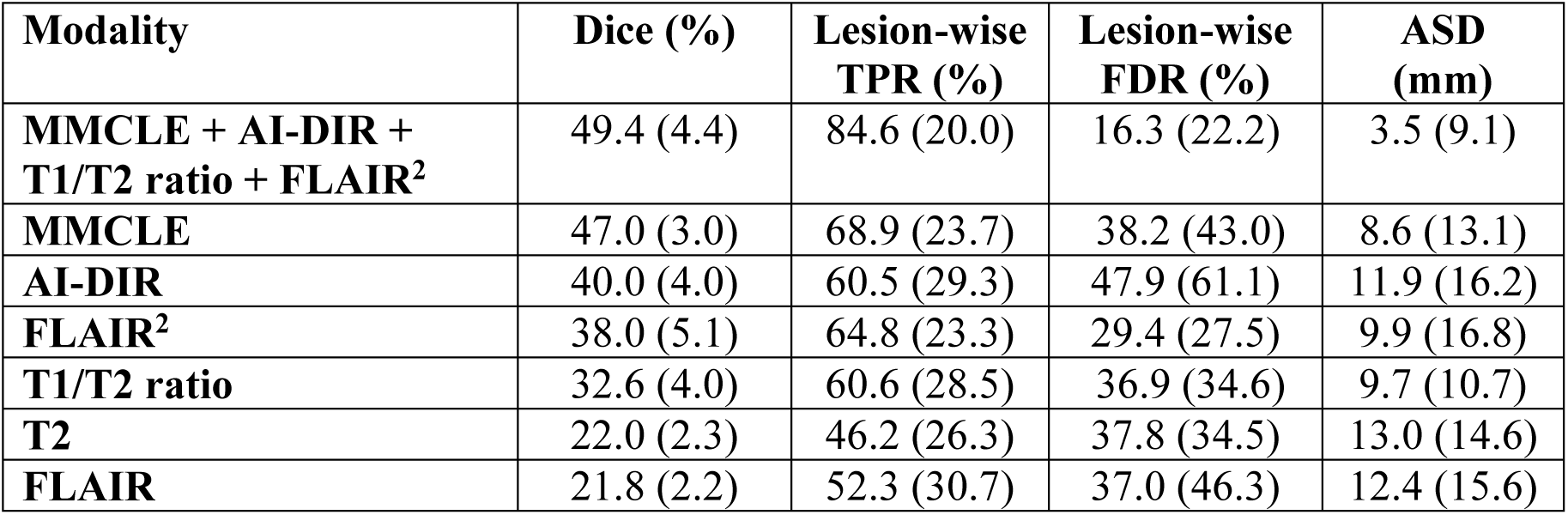
AI training results on the 80 selected patients, presented as mean (standard deviation). Dice represents Sørensen–Dice index. Lesion-wise true positive rate is the proportion of true lesions correctly classified by the algorithm. Lesion-wise false discovery rate (FDR) is the proportion of algorithm-proposed lesions that are not true lesions. MMCLE – multi-modal cortical lesion enhanced; AI-DIR – artificial intelligence double inversion recovery; TPR – true positive rate; ASD – average surface distance metric. Rows are listed in descending order of Dice.

For each lesion, we also identified whether or not it was independently visible on each of the 6 contrast types and tabulated the relative proportion of lesions having each of the 2^6^=64 potential combinations of visibility (Supplementary Table 1). The most common pattern(18.2% of lesions) was visibility on MMCLE, AI-DIR, FLAIR^2^, T1/T2 ratio, and T2, but not on FLAIR. The next highest pattern(13.2% of lesions) was detectability on all four synthetic contrasts (MMCLE, AI-DIR, FLAIR^2^, and T1/T2 ratio), but not on conventional FLAIR or T2. Another 7.2% of lesions were seen on MMCLE, AI-DIR, and T1/T2 ratio, but not FLAIR^2^, FLAIR, or T2, and 7.1% were seen on MMCLE and T1/T2 ratio only. The remaining lesions were seen on a variety of combinations, except for 3.8% that were originally detected when viewing all contrasts simultaneously but were not seen in a blinded manner on any independent contrast alone. Notably, 9.7% of lesions were seen on only a single contrast, with 4.7% visible on MMCLE only, 3.9% on T1/T2 ratio only, 0.7% on AI-DIR only, and 0.5% on FLAIR^2^ only. No lesions were seen solely on conventional FLAIR or T2 contrasts. Conversely, 5.1% of lesions were missed on MMCLE but seen on at least one other contrast. This rate was 9.1% for AI-DIR, 5.9% for T1/T2 ratio, and 9.2% for FLAIR^2^.

### AI training underscores the need for simultaneous review of multiple contrasts

Validation metrics for AI training on the 80 analyzed cases are reported in Table 3. Dice metrics were significantly different based on a Kruskal-Wallis test (p<0.001), with the highest values for the combined training on all contrasts, and lowest for the T1/T2 ratio alone.

For the final deployment AI lesion detection model trained on the full dataset on all cortical-lesion-sensitive contrasts, Dice was 59.9 (SD=1.3). For the model trained on the full dataset with raw images (T1, T2, FLAIR), Dice was 53.1 (SD=1.5). On the model trained on the longitudinal full dataset, Dice for new lesions was 15.6 (SD=4.3) and Dice for enlarging lesions was 8.4 (SD=6.1).

## Discussion

In this study, we evaluated the potential benefit of different post-processing and synthetic contrast generation techniques for post-hoc detection of cortical lesions in MS clinical trial datasets that were acquired with conventional sequences. We found that, indeed, these techniques can be directly and meaningfully applied to these datasets to reveal many cortical lesions that would otherwise remain invisible. We also found that the benefit of this reprocessing is maximized when the different techniques are employed together due to their different strengths, particularly when combined via MMCLE. Our overall findings are also consistent with prior studies on the individual contrasts.^28,29,33^ We used three different approaches to support our findings: quantitative measurements of CNR; blinded, sham-based rater reviews; and assessment of the ability of deep learning to accurately quantify lesions. In all cases, the MMCLE approach (combining AI-DIR and FLAIR^2^) provided the best detectability compared to simultaneous review of all contrasts. In particular, MMCLE showed a more than 2.3-fold increase in CNR compared to conventional imaging (T2-weighted and FLAIR) for distinguishing lesions from normal-appearing gray matter. This increased rater detectability by 2.3-fold, and improved AI detectability by 32%. Furthermore, the use of multiple simultaneous contrasts showed a clear advantage, with 3.8% of lesions seen only when multiple contrasts were available. Similarly, using all four synthetic contrasts improved AI detectability by 1.62-fold compared to conventional imaging and provided substantially improved false positive rate (FPR) and true positive rate (TPR) results compared to any individual contrast.

Given the prevalence of cortical lesions in MS, their clinical and cognitive relevance, and their potentially direct pathophysiological involvement with gray matter damage, our confirmation of multi-contrast cortical lesion detectability on conventionally acquired clinical trial data is highly encouraging. It indicates that we can re-use large, costly, carefully planned, blinded, experimentally controlled datasets to better understand the relationship of cortical lesions to MS diagnosis, prognosis, and treatment response, and to better understand the impact of MS treatments and drug classes on this unique pathology.^19^ It also means that when practical constraints prevent the inclusion of advanced sequences (such as DIR) in scanning protocols in future studies, it is still possible to include cortical lesions as outcome measures even with the typical sequences routinely used for subcortical lesion detection. With further research, it may also improve McDonald-criteria-based diagnosis by making cortical lesion detection more attainable in clinical practice.

All of the contrasts used here are relatively straightforward to implement. FLAIR^2^ and T1/T2 ratio are simple multiplicative combinations than can be produced easily after basic and widely-used pre-processing steps, including co-registration and intensity adjustment. The AI-DIR contrast is conceptually complex, but is also easy to implement in practice on any modern GPU. MMCLE, like FLAIR^2^ and T1/T2 ratio, is also based on simple voxel-wise multiplication. This means that these approaches are not only possible, but also highly feasible.

During the study, we also made some specific observations about each of the contrasts of interest. In particular, and consistently with the quantitative results, lesions were most salient on MMCLE, standing out as clear areas of hyperintensity. AI-DIR was notably smoothest, likely resulting from the regularizing influence of the generative model. As an AI-based approach, there is also a natural concern for potential hallucination – the artificial creation of data based on training expectations rather than on the actual data from the current case.^34^ Therefore, *a priori*, we were concerned about the possibility of AI-DIR introducing spurious lesions. However, we did not find examples of this, and instead found occasional “negative hallucinations” where the AI-DIR “filled in” lesions that were more clearly visible on the conventional images (e.g. rows 3 and 5 in Figure 1 show less contrast than conventional images). In contrast, FLAIR^2^ tended to be noisier, but showed most lesions (albeit with less salience than MMCLE). Its direct relationship to truly acquired images also improved confidence in classifications. Finally, T1/T2 ratio appeared to confirm nearly all lesions when viewed *a posteriori*. However, because both cortical lesions and many other features are hypointense on T1/T2 ratio, they do not stand out nearly as much as on other sequences. As such we found it to be best used as a confirmatory sequence rather than a primary sequence for detection. While we did not undertake a separate study to quantify it, we also noticed that lesions appeared visually somewhat larger on T1/T2 ratio than on other contrasts. This should likely be further investigated, and should be considered when comparing studies using different cortical-lesion-sensitive methods. It would also be important to systematically explore why some lesions are seen on specific sequences and not others.

While the value of cortical lesion data in better characterizing MS pathological status and progression argues strongly for its assessment using these techniques, there are some areas of caution that should be carefully considered. One concern (which applies to all current imaging methods) is detection bias for certain subtypes of cortical lesion. We know from pathological data that subpial lesions are the most prominent type by volume,^9^ but these lesions are the most poorly resolved type by in vivo MRI, even at 7T and with bespoke cortical-lesion-sensitive contrasts.^22,26^ Although we did not explicitly classify the lesion types in this study, we observed a similar pattern, with lesions appearing more salient when they included white matter involvement. Because they remain largely driven by T2 effects, there is no reason to expect that the post-processed contrasts used here would improve the visibility of subcortical lesions beyond the current state of the art imaging methods. Fortunately, prior research has shown that the fraction of 3T-visible subpial lesions is highly correlated with total 7T-visible volume.^23^ Nonetheless, future studies should carefully consider the likely underestimation of subpial lesion presence and/or activity, and should also continue to search for new MRI contrasts more sensitive to these lesions in the challenging low-myelin subpial area.

The impact of study-to-study protocol differences also merits consideration, since the AI-DIR method was initially trained primarily on images with 3D acquisitions, and FLAIR^2^ early validation was based on 3D-FLAIR. However, we worked on a broad set of protocols from a variety of scanners in this study. We also experimented on an internal dataset for which we had both 2D T1 spin echo and 3D T1 GRE, and found ICC>0.96 (absolute agreement) for cortical lesion counts and volumes. Nonetheless, this should be more systematically studied.

Another potential issue is that cortical lesion scoring remains relatively difficult compared to traditional white matter lesions, even on acquisitions prospectively tailored for it, including true DIR. In previous multi-center work, majority agreement was found for only 54.0% of lesions and full agreement across 5 raters was only found for 19.4% of lesions.^24^ AI tools have the potential to help obviate some of these issues by providing consistency, as demonstrated on cortical-lesion-sensitive contrasts by prior studies.^35^ Our work here indicates that this is also possible on post-processed contrasts like FLAIR^2^, AI-DIR, T1/T2 ratio and MMCLE, resulting in Dice scores in line or better than previously reported inter-rater measures.^35^ However, as with any AI tool, our classifier may be limited by the training data quality and generalizability.

In addition to these broader concerns, there are some limitations that are specific to our study and that should be considered when generalizing our results. A principal limitation is that we had no reference standard against which to compare the true sensitivity and specificity of these contrasts. Histopathology would have provided valuable additional data. Similarly, 7T or advanced, cortical-lesion-sensitive 3T sequences could have served as a surrogate, but were not available for this retrospective dataset. Although our results are in line with previous reports,^22,23,36,37^ it would nonetheless be important in future work to evaluate sensitivity and specificity via a reference standard. Further work is under way to explore clinical validity of these measures and treatment effects in the full ORATORIO dataset. Another study-specific limitation is that the population we studied consisted entirely of PPMS. Although cortical lesions have been shown to be present in comparable numbers in progressive MS,^20^ and although this was mainly an investigation of the characteristics of image contrasts rather than disease characteristics, it would still be helpful to further study our approach in other MS populations. In particular, use of PPMS may bias the relative findings against the more sophisticated AI-DIR technique that has been trained on specific underlying distributions, and may not reflect its true potential if re-trained on a tuned dataset. Finally, while we had a wealth of cross-sectional cortical lesion data to train AI tools on, longitudinal cortical lesion activity was very sparse. In contrast to the excellent Dice metrics for cross-sectional lesion detection and delineation, this resulted in poor Dice measures for our activity classifier. Future work with a larger and/or more active dataset should seek to train better longitudinal AI models to help improve the process of activity assessment for cortical lesions.

In conclusion, there is a substantial capacity for the combined use of these approaches to quantify cortical lesion burden and evolution in MS, allowing us to re-use and re-evaluate extensive clinical trial datasets to understand cortical lesion evolution and treatment with experimental data, as well as add flexibility in the design of future studies by offering an alternative to the inclusion of advanced sequences when cortical lesions are a desirable outcome measure. Given the importance of these lesions, this should be urgently investigated in the many already-available clinical-trial datasets across different therapeutic mechanisms.

## Methods

### Study design and participants

This was a retrospective analysis of a subset of data from the multicenter, Phase III ORATORIO study in which 732 subjects with primary progressive MS were randomized to ocrelizumab or placebo.^31^ Because the goal of the current study was to establish and evaluate methodology, and not to determine treatment effect (which will be part of future work), a total of 80 subjects were randomly selected from this pool, stratified by total T2 lesion volume (8 subjects randomly selected from each decile of T2 lesion volume). This subset was used for all development and analysis except for a single, final training of a full AI model for future use on all scans.

### Image acquisition and pre-processing

Brain MRI images were acquired in the ORATORIO study at screening (baseline) and weeks 24, 48, and 120. The following sequences were obtained using 3-mm axial slices (no gap) and whole brain coverage and 1×1 mm^2^ in-plane resolution: proton density and T2-weighted two-dimensional (2D) multislice turbo/fast spin-echo; T1-weighted three-dimensional (3D) spoiled gradient recalled echo (pre-contrast and post-contrast); and 2D T2-weighted Fluid-Attenuated Inversion Recovery (FLAIR, post-contrast).

Additional acquisition information is provided in the supplementary material to the original study manuscript.^31^ All raw images were processed with N4 to correct for bias field inhomogeneities.^38^ Subsequently, baseline T1-weighted pre-contrast images (T1s) were aligned to MNI space^39^ using 12 degrees of freedom, and the rigid portion of the obtained matrix (6-dof) was used to provide a roughly aligned but volume-preserved image space using FSL FLIRT.^40^ Other baseline contrasts and all follow-up images were registered to the baseline MNI-aligned T1s using six degrees of freedom, and all images were upsampled to 1mm isotropic voxels using spline interpolation. BET was employed on T1s to perform deskulling,^41^ and conventional T2 lesions were inpainted using FSL’s lesion_filling tool.^40^ To provide additional context during manual lesion delineation, FastSurfer was used to generate cortical surface maps.^42^ These maps were augmented by including a 2mm dilation at a lower intensity to highlight areas within 2mm of the cortex, as well as by removing any voxels within 2mm of the lateral ventricles. Finally, although not directly used for cortical lesion detection, we also performed tissue segmentation with FSL’s SIENAX and conventional T2 lesion detection using an in-house deep-learning system with manual review^43^ to produce normal-appearing tissue maps for contrast-to-noise (CNR) measurements.^44^ Figure 2 shows a schematic overview of the proposed pipeline.

**Figure 2.**
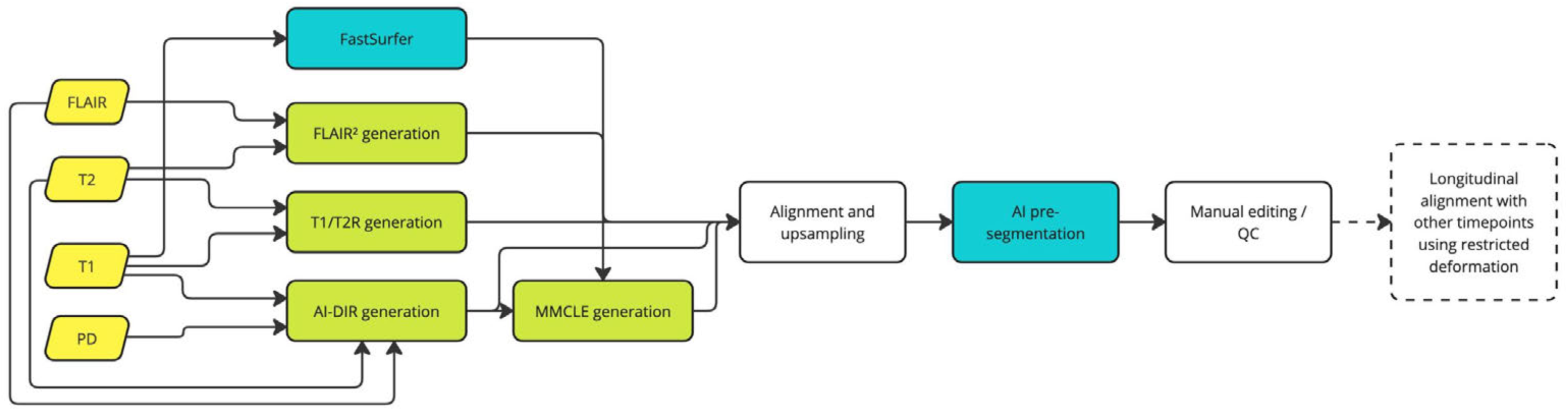
Overview of the proposed pipeline for cortical lesion analysis on legacy images. Conventional FLAIR, T2, T1, and PD images are used to produce four cortical-lesion-sensitive contrasts. These are all aligned and upsampled to a unified 1mm isotropic space, pre-segmented via AI, and then presented to a user via a custom Slicer module for manual editing and review. Restricted-deformation warping is used to provide longitudinal subtractions.

### FLAIR^2^ creation

FLAIR^2^ images were created as described by Zrzavy et al.,^33^ based on multiplication of aligned FLAIR and T2 images. Because image intensities were scaled and offset differently between centers and acquisitions, input images were first intensity standardized based on the 25^th^ and 99.75^th^ percentiles of intensities within the BET-derived brain mask.

### T1/T2 ratio creation

T1/T2 ratio images were created as described by Manning et al.,^28^ by dividing T1 images by the T2 images. Because T2 images were available, we used these rather than FLAIR images, to avoid division by values close to zero in areas of CSF suppression. Additionally, as for FLAIR^2^ creation, intensities were first standardized based on 25^th^ and 99.75^th^ percentiles within the brain mask region.

### AI-DIR creation

AI-DIR images were created as described by Bouman et al.^29,30^ Briefly, aligned individual images were z-score-adjusted and then fed into a fully convolutional U-shaped (encoder/decoder) generator that was previously trained using a generator/discriminator-based generative adversarial network (GAN) approach. For this implementation, all four conventional contrasts – T1 pre-contrast, T2, FLAIR, and PD – were employed.

### MMCLE creation

In preliminary experiments, we noted visually complementary information in AI-DIR and FLAIR^2^ images. To exploit this, we produced a combination of the two via voxel-wise multiplication with equal weighting for each input image (extending the core idea underlying FLAIR^2^). A representative example of this combination is shown in Figure 3.

**Figure 3.**
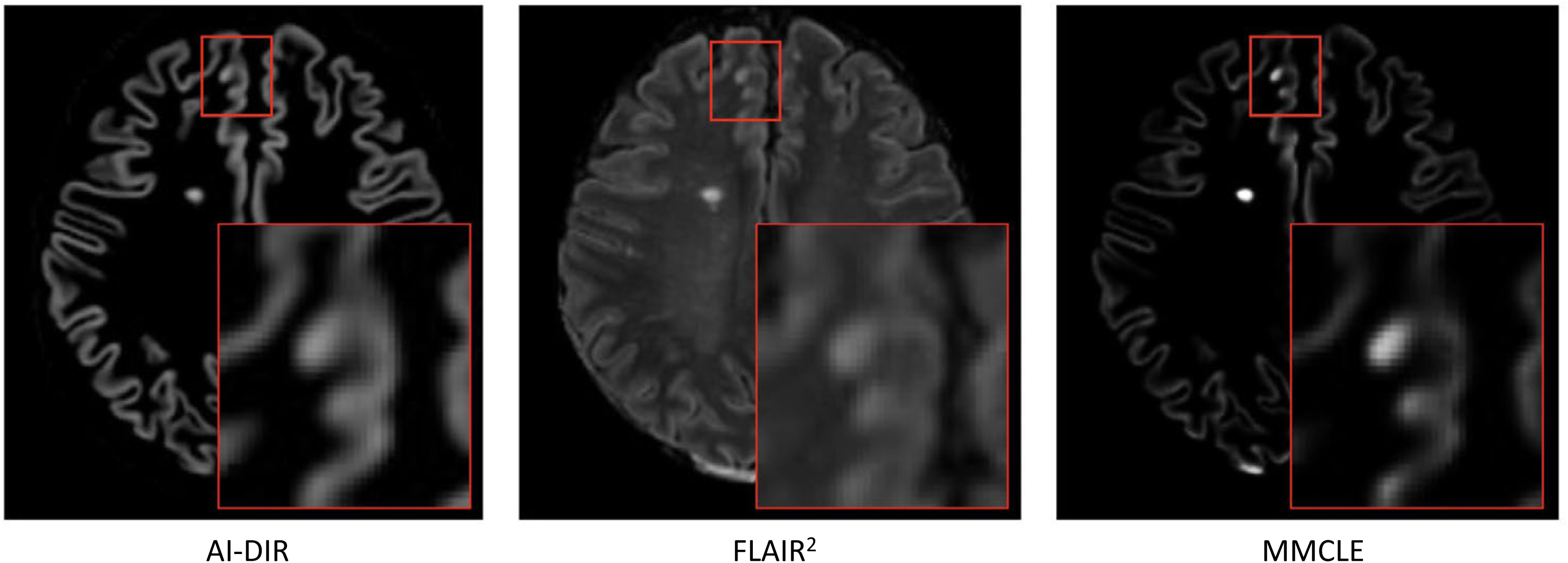
Representative example of the creation of a multi-modal cortical lesion enhanced (MMCLE) image from input artificial-intelligence double inversion recovery (AI-DIR, left) and fluid attenuated inversion recovery squared (FLAIR^2^, center) images. The MMCLE approach substantially improves the contrast to noise, and therefore saliency, of cortical lesions, as confirmed in Table 2.

### Follow-up warping and subtraction imaging

While broadly fixed, the cortex may undergo small local shifts due to atrophy or biological fluctuations over time. Because these changes are local and/or nonlinear, they are not fully addressed by linear registration. Although often negligible in white matter subtraction imaging, we found that, due to the thinness of the cortex, these small residual displacements substantially impacted the quality of longitudinal subtraction. Therefore, we employed an additional non-linear warping step between baseline and follow-up images using ANTs.***^45^***Critically, we limited the scale and iterations to only allow for relatively coarse displacements (400x100x50 iterations at 8x4x2 shrink factors) in order to minimize impact on the cortical lesions themselves. We then performed conventional subtraction imaging on intensity-matched, warped images.***^46^***Figure 4 illustrates the impact of this additional warping step.

**Figure 4.**
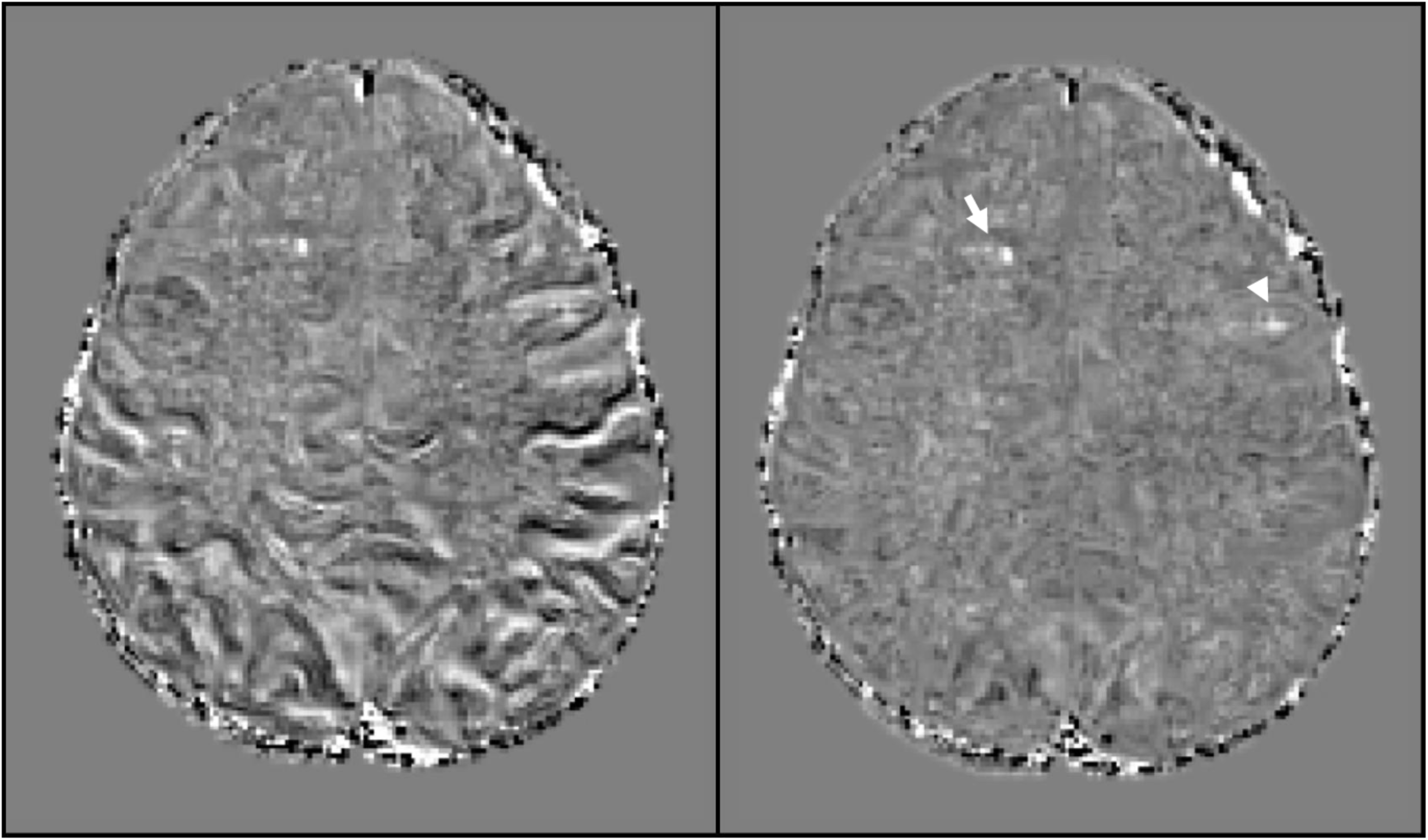
Residual non-linear displacements due to atrophy, biological fluctuation, and uncorrected distortions may impact cortical lesion detection more substantially than white matter lesion detection, but may be further corrected by non-linear longitudinal warping, as shown here. On the left is a T1/T2 ratio subtraction image based on linear warping alone, and on the right is the same image with the inclusion of additional warping between timepoints. Contrast is inverted for visualization, so that new lesions are shown as white areas, with a new white matter lesion that is clear on both images (arrow) and a new cortical lesion that is easily missed on the non-warped left image (arrowhead).

### Primary lesion analysis

To facilitate lesion analysis, a custom 3D Slicer^47^ module was created to simultaneously view all relevant images, including MMCLE, FLAIR^2^, cortex map, T1/T2 ratio, AI-DIR, and FLAIR. At baseline, the MMCLE, FLAIR^2^, cortex map, T1/T2 ratio, AI-DIR, and FLAIR images were used, as shown in Figure 5. At follow-up, paired baseline and follow-up MMCLE, T1/T2 ratio, and FLAIR^2^ images, follow-up cortex map, subtraction images, and a non-warped baseline MMCLE (to confirm lack of warping interference on cortical lesion size) were used, as shown in Figure 6.

**Figure 5.**
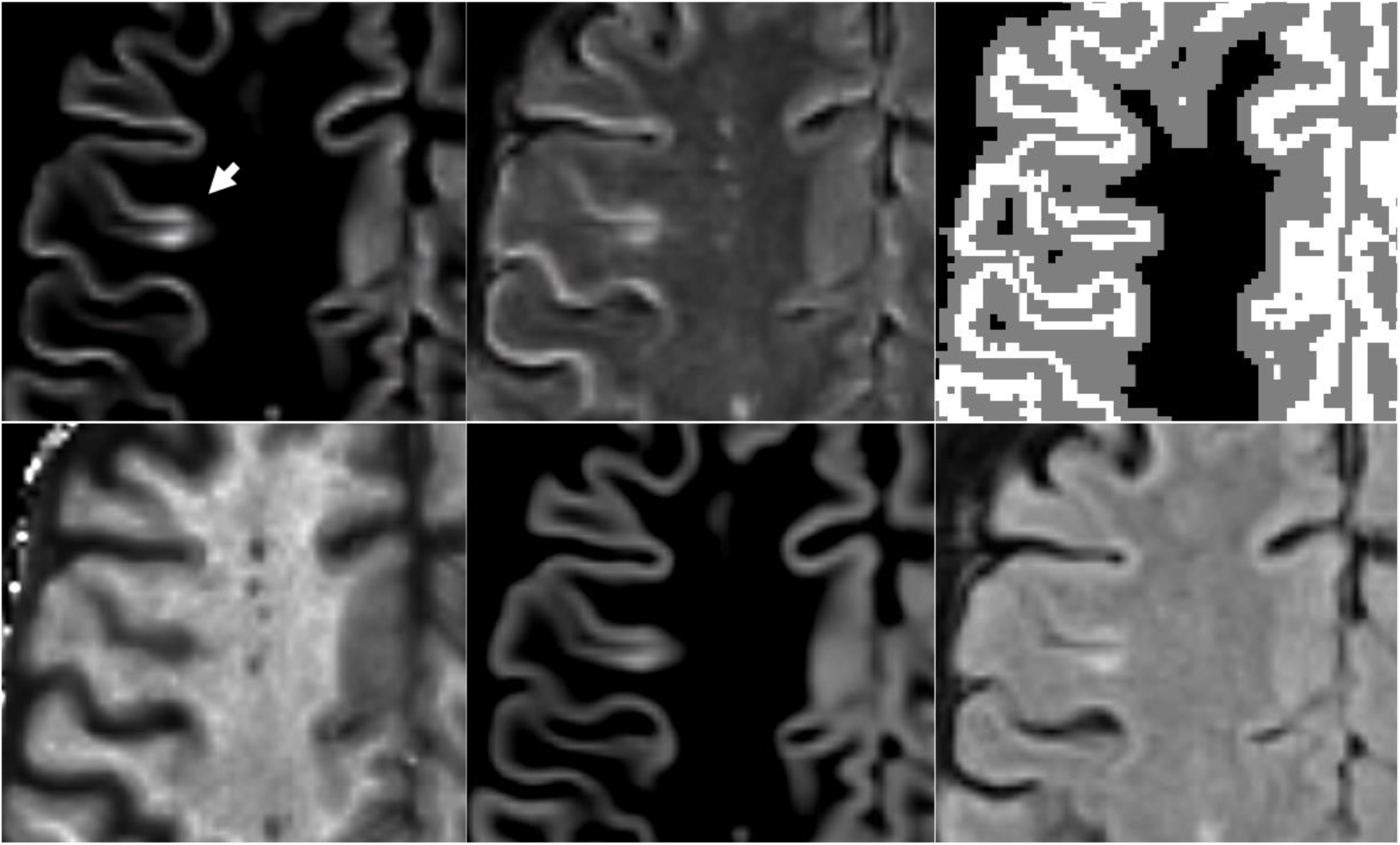
Representative example of the images and layout used for simultaneous for lesion detection and/or review. An example of a well-visualized subpial lesion is highlighted (arrow). Upper left: multi-modal cortical lesion enhanced (MMCLE) image, a combination of FLAIR^2^ and AI-DIR. Upper middle: FLAIR^2^, a combination of FLAIR and T2. Upper right: cortical map from FastSurfer (actual cortex shown in white, voxels within 2mm of cortex, including from above and below, in gray). Lower left: T1/T2 ratio image. Lower middle: AI-DIR, synthesized based on a deep neural network from the FLAIR, T1, PD, and T2 images. Lower right: conventional FLAIR.

**Figure 6.**
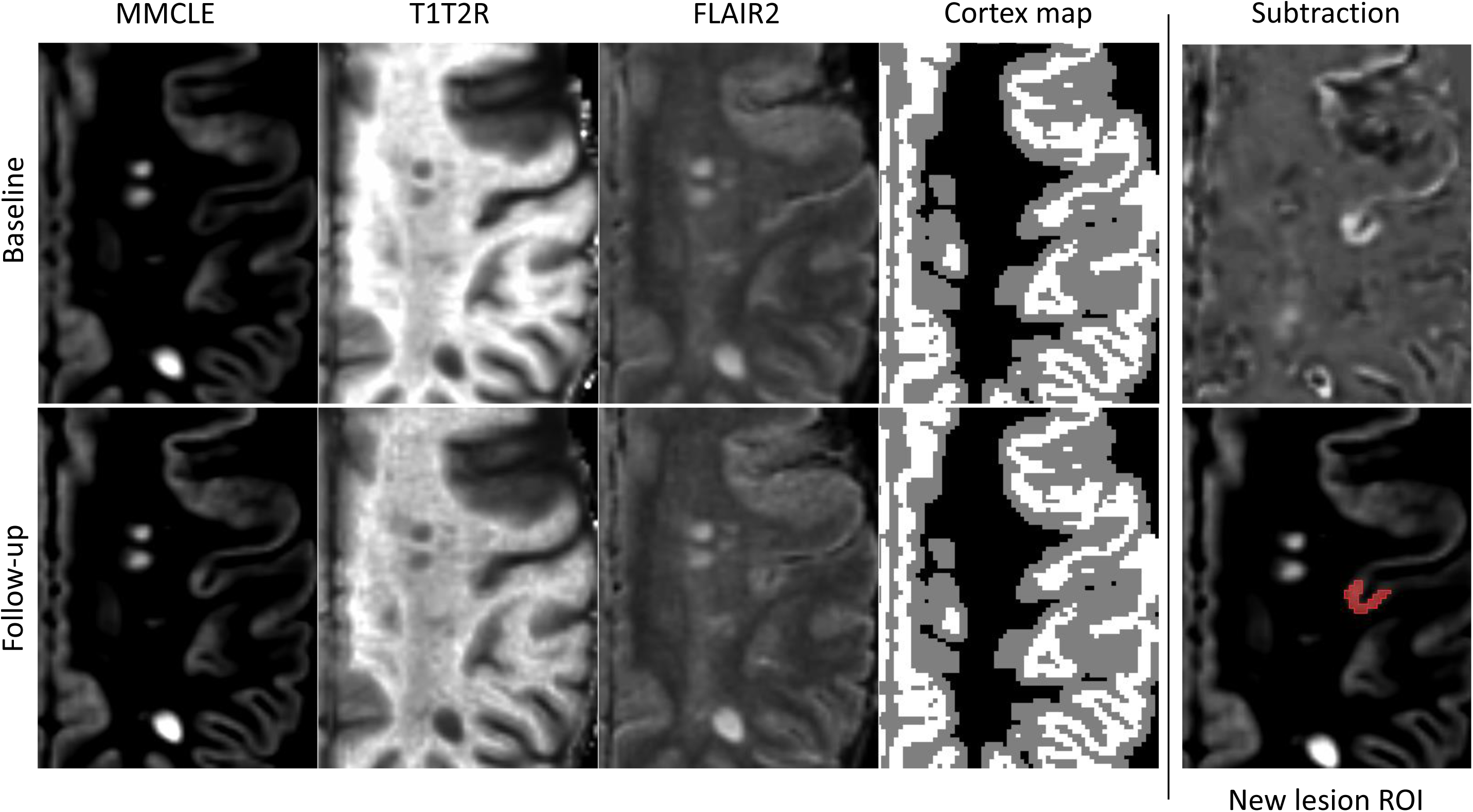
Visualization of follow-up analysis process showing paired baseline (upper) and follow-up (lower) MMCLE, T1/T2 ratio, and FLAIR^2^ images, follow-up cortex map, subtraction image (upper right), and a non-warped baseline MMCLE (to confirm lack of warping interference on cortical lesion size) with ROI overlaid (lower right).

Cortical lesions were manually labelled by four trained raters (MD, NB, DJ, AB) with 21, 20, 9, and 7 years of MS imaging experience. All cases were then reviewed by an expert rater (RZ) with 25 years of MS imaging experience and consensus was reached. Lesions were identified in line with pre-defined criteria.^24^ In particular, they were required to be brighter than normal gray matter on FLAIR^2^, AIDIR, and MMCLE, or darker than normal gray matter on T1/T2 ratio. They were also required to be at least 3 voxels in size, and to not be commonly seen artifacts or traceable cortical vessels. To help distinguish between u-fiber lesions and true cortical involvement, we additionally required all lesions to have at least 2 voxels fully within the cortex or directly touching the cortical surface. In the case of confluent lesions, only the portions of lesions within the 2mm neighborhood of cortex were included, and only when the cortical lesion could be clearly visually separated from the rest of the confluent complex. At follow-up, new and enlarging lesions were required to be areas of clear hyperintensity on MMCLE or FLAIR^2^ subtraction, or of clear hypointensity on T1/T2 ratio subtraction, and enlarging lesions were required to increase in volume by at least 50%. Finally, to avoid potential AI-based hallucination, lesions were required to be visualized on at least two contrasts, one of which was required to be non-AI (FLAIR^2^ or the T1/T2 ratio). This primary, multi-contrast lesion analysis was used as the reference standard for subsequent analyses of individual contrasts.

### Lesion contrast characteristics

To better understand the characteristics of individual sequences, we assessed lesion-wise CNR as compared to the normal-appearing tissue, as well as to the normal-appearing gray matter and white matter separately. To identify normal appearing tissue, cortical lesions masks were combined with conventional T2 lesion masks, dilated, inverted, and multiplied by the relevant tissue maps from SIENAX. CNR measures were calculated within each subject and then averaged across all subjects.

### Blinded per-contrast analysis

Because we sought to compare the relative visibility of lesions on each contrast, we also conducted a post-hoc, randomized, blinded review of all lesions. For each previously identified lesion on each cortical-lesion-sensitive contrast, a square ROI extending 2 cm from the lesion on all sides was cropped and saved as a separate image. At the same time, for each lesion, a “sham” region was selected from the same subject’s image set such that it contained no central lesion but was the same distance from the cortex as the true lesion (including within the cortex). For comparison, we also included conventional T2 and FLAIR images. This resulted in 12 images per lesion – one real for each contrast and one sham for each contrast. Then, all lesions across all subjects and all contrasts were randomized and presented one at a time via a web interface developed in Flask (version 3.1.0, 2024) to three independent reviewers (MD, NB, AB) who classified each as either real or sham. All responses were recorded for later statistical analysis. To minimize recall bias, this analysis was performed more than 3 months after the original analysis.

### AI training and multi-contrast evaluation

In order to evaluate the relative contributions of the various cortical-lesion-sensitive MRI contrasts, we trained a set of deep learning artificial neural networks to recognize and delineate cortical lesions on each of the contrasts individually and on them all simultaneously, as well as on conventional T2 and FLAIR for comparison.

To take advantage of recent advances in deep learning architecture, we used a sliding window (Swin) UNEt Transformer (Swin-UNETR)^48^ rather than a conventional Unet architecture. Swin-UNETR combines the strengths of convolutional neural networks’ (CNN) hierarchical feature learning with transformer-style self-attention^49^ by using shifted window (Swin) transformers as encoders in a U-shaped network connected to a multiresolution CNN-based decoder via skip connections. To overcome the greater training data requirements of transformer-based architectures, we initialized the model with weights from a publicly available pre-training run.^50^ Data augmentations including random cropping (balanced between classes), random flips and rotations, and random intensity scaling and shifts were employed. Training then proceeded for 100 epochs with 1 warm-up epoch and early stopping, using AdamW^51^ as the optimizer and an equally-weighted combination of Dice and cross-entropy (CE) as the loss function.^52^ A learning rate of 0.0004 was employed with linear warmup and cosine decay. All code was implemented via the medical open network for AI (MONAI) framework (version 1.3.0 2023) and PyTorch (version 2.0.0 2023). The same architecture was used to train individual models for each contrast or combination of contrasts, with the only modification being the number of channels for the initial input. All training was performed on a system with 48 Intel Xeon CPUs, 192 GB RAM, and 4 Nvidia T4 GPUs with a total of 64GB GPU memory.

In addition to these models, which were each trained on the 80-subject baseline dataset, three further models were trained on the full complement of 732 subjects’ data to provide an objective, rapid, and reliable classification procedure for future analyses. This full training set was bootstrapped by using the 4-modality, 80-subject classifier to produce initial cortical lesion masks, which were then manually reviewed and corrected as in the primary lesion analysis. These models used the same architecture as the prior models, only differing in the number of input channels. First, to provide the most comprehensively trained model possible, a network was trained on all four cortical-lesion-sensitive contrasts (AI-DIR, MMCLE, T1/T2 ratio and FLAIR^2^) on all baseline scans. Second, based on the consideration that the post-processed cortical-lesion-sensitive contrasts are intermediate products from the raw images, and that the pattern-recognition capacity for deep learning may obviate the need for these intermediate steps and be able to directly identify cortical lesions on raw images, we trained a model on T1 pre-contrast, T2, FLAIR, and PD images alone. Finally, to assist with longitudinal lesion classification, another model was trained on all four cortical-lesion-sensitive contrasts across two timepoints to detect new and enlarging activity.

### Statistical analysis

For the blinded per-contrast analysis (described above), agreement between the three raters was assessed via Fleiss’ kappa. Individual raters’ agreements with ground truth were assessed via Cohen’s kappa as well as true positive rate (TPR) and false positive rate (FPR). Data was also analyzed on a per-lesion basis to determine the proportion of cortical lesions seen by the majority (2/3) of raters on specific combinations of contrasts. Analyses were performed in R (version 4.4.0, R Core Team 2024) using the irr package (version 0.84.1 2019). For AI training, five-fold cross-validation was used to evaluate the accuracy and variability of training on each image type or combination, with Dice score as the primary validation criterion. Additional lesion-wise statistics included true positive rate (TPR), false discovery rate (FDR), and the average surface distance (ASD).^53^ Calculations were performed in Python (3.10.11). All lesion statistics were calculated first at the subject level and then averaged across subjects, with the exception of the per-lesion data reported in Supplementary Table 1.

## Data Availability

All data produced in the present study are available upon reasonable request to the authors.

## Supplemental figure

**Table S1.** Prevalence of each combination of lesion visibilities, as assessed by sham-blinded, randomized review of each lesion on each contrast. Each row is a unique combination of visibilities.

| Contrast type |  |  |  |  |  | Frequency |  |
| --- | --- | --- | --- | --- | --- | --- | --- |
| MMCLE | AI-DIR | FLAIR<sup>2</sup> | T1/T2 ratio | FLAIR | T2 | % | Cumulative % |
| Yes | Yes | Yes | Yes | No | Yes | 18.19 | 18.19 |
| Yes | Yes | Yes | Yes | No | No | 13.2 | 31.39 |
| Yes | Yes | No | Yes | No | No | 7.19 | 38.58 |
| Yes | No | No | Yes | No | No | 7.11 | 45.69 |
| Yes | No | Yes | Yes | No | No | 5.75 | 51.44 |
| Yes | No | Yes | Yes | No | Yes | 5.67 | 57.11 |
| Yes | Yes | Yes | Yes | Yes | Yes | 5.67 | 62.77 |
| Yes | No | No | No | No | No | 4.65 | 67.43 |
| No | No | No | Yes | No | No | 3.89 | 71.32 |
| Yes | Yes | Yes | Yes | Yes | No | 3.89 | 75.21 |
| No | No | No | No | No | No | 3.81 | 79.02 |
| Yes | Yes | No | No | No | No | 3.72 | 82.74 |
| Yes | Yes | Yes | No | No | No | 3.55 | 86.29 |
| Yes | Yes | No | Yes | No | Yes | 1.78 | 88.07 |
| Yes | No | No | Yes | No | Yes | 1.44 | 89.51 |
| Yes | No | Yes | No | No | No | 1.27 | 90.78 |
| Yes | No | Yes | No | No | Yes | 0.93 | 91.71 |
| No | No | No | Yes | No | Yes | 0.85 | 92.55 |
| No | Yes | No | No | No | No | 0.68 | 93.23 |
| Yes | Yes | Yes | No | Yes | No | 0.68 | 93.91 |
| No | No | Yes | Yes | No | No | 0.68 | 94.59 |
| Yes | No | Yes | Yes | Yes | No | 0.68 | 95.26 |
| Yes | Yes | No | Yes | Yes | No | 0.59 | 95.85 |
| Yes | Yes | Yes | No | No | Yes | 0.59 | 96.45 |
| No | No | Yes | No | No | No | 0.51 | 96.95 |
| Yes | No | Yes | Yes | Yes | Yes | 0.42 | 97.38 |
| No | Yes | No | Yes | No | No | 0.34 | 97.72 |
| No | No | Yes | Yes | No | Yes | 0.34 | 98.05 |
| Yes | No | Yes | No | Yes | No | 0.25 | 98.31 |
| Yes | Yes | No | No | Yes | No | 0.17 | 98.48 |
| No | Yes | Yes | Yes | No | No | 0.17 | 98.65 |
| Yes | Yes | Yes | No | Yes | Yes | 0.17 | 98.82 |
| No | Yes | No | Yes | No | Yes | 0.17 | 98.98 |
| No | No | Yes | Yes | Yes | Yes | 0.17 | 99.15 |
| No | No | Yes | No | Yes | No | 0.08 | 99.24 |
| Yes | No | No | No | Yes | No | 0.08 | 99.32 |
| No | No | Yes | Yes | Yes | No | 0.08 | 99.41 |
| No | Yes | Yes | Yes | Yes | No | 0.08 | 99.49 |
| Yes | No | No | Yes | Yes | No | 0.08 | 99.58 |
| No | No | Yes | No | No | Yes | 0.08 | 99.66 |
| Yes | No | No | No | No | Yes | 0.08 | 99.75 |
| Yes | No | Yes | No | Yes | Yes | 0.08 | 99.83 |
| No | Yes | Yes | Yes | No | Yes | 0.08 | 99.92 |
| Yes | Yes | No | Yes | Yes | Yes | 0.08 | 100 |
| No | No | No | No | Yes | No | 0 | 100 |
| No | Yes | No | No | Yes | No | 0 | 100 |
| No | Yes | Yes | No | No | No | 0 | 100 |
| No | Yes | Yes | No | Yes | No | 0 | 100 |
| No | No | No | Yes | Yes | No | 0 | 100 |
| No | Yes | No | Yes | Yes | No | 0 | 100 |
| No | No | No | No | No | Yes | 0 | 100 |
| No | Yes | No | No | No | Yes | 0 | 100 |
| No | No | No | No | Yes | Yes | 0 | 100 |
| No | Yes | No | No | Yes | Yes | 0 | 100 |
| No | Yes | Yes | No | No | Yes | 0 | 100 |
| No | No | Yes | No | Yes | Yes | 0 | 100 |
| No | Yes | Yes | No | Yes | Yes | 0 | 100 |
| Yes | Yes | No | No | No | Yes | 0 | 100 |
| Yes | No | No | No | Yes | Yes | 0 | 100 |
| Yes | Yes | No | No | Yes | Yes | 0 | 100 |
| No | No | No | Yes | Yes | Yes | 0 | 100 |
| No | Yes | No | Yes | Yes | Yes | 0 | 100 |
| No | Yes | Yes | Yes | Yes | Yes | 0 | 100 |
| Yes | No | No | Yes | Yes | Yes | 0 | 100 |

